# Protocol for a Delphi consensus process for PARticipatory Queer AI Research in Mental Health (PARQAIR-MH)

**DOI:** 10.1101/2023.08.07.23293764

**Authors:** Dan W Joyce, Andrey Kormilitzin, Julia Hamer-Hunt, Kevin R McKee, Nenad Tomasev

**Affiliations:** Department of Primary Care and Mental Health, Institute of Population Health, University of Liverpool, United Kingdom, L69 3GF; Department of Psychiatry, University of Oxford, Warneford Hospital, Oxford, United Kingdom, OX3 7JX; Google DeepMind, London, United Kingdom

## Abstract

**Introduction:** For artificial intelligence (AI) to help improve mental health care, the design of data-driven technologies needs to be fair, safe, and inclusive. Participatory design can play a critical role in empowering marginalised communities to take an active role in constructing research agendas and outputs. Given the unmet needs of the LGBTQI+ community in mental health care, there is a pressing need for participatory research to include a range of diverse queer perspectives on issues of data collection and use (in routine clinical care as well as for research) as well as AI design. Here we propose a protocol for a Delphi consensus process for the development of PARticipatory Queer AI Research for Mental Health (PARQAIR-MH) practices, aimed at informing digital health practices and policy.

**Methods and Analysis:** The development of PARQAIR-MH is comprised of four stages; In Stage 1, a review of recent literature and fact-finding consultation with stakeholder organisations will be conducted to define a terms-of-reference for Stage 2, the Delphi process. Our Delphi process consists of three rounds, where the first two rounds will iterate and identify items to be included in the final Delphi survey for consensus ratings. Stage 3 consists of consensus meetings to review and aggregate the Delphi survey responses leading to Stage 4 where we will produce a reusable toolkit to facilitate participatory development of future bespoke LGBTQI+–adapted data collection, harmonisation and use for data-driven AI applications specifically in mental health care settings.

**Ethics and Dissemination:** The PARQAIR-MH aims to deliver a toolkit that will help to ensure that the specific needs of LGBTQI+ communities are accounted for in mental health applications of data-driven technologies. Participants in the Delphi process will be recruited by snowball and opportunistic sampling via professional networks and social media (but not by direct approach to healthcare service users, patients, specific clinical services or via clinicians’ caseloads). Participants will not be required to share personal narratives and experiences of healthcare or treatment for any condition. Before agreeing to participate, people will be given information about the issues considered to be in-scope for the Delphi (e.g. developing best practices and methods for collecting and harmonizing sensitive characteristics data; developing guidelines for data use/re-use) alongside specific risks of unintended harm from participating that can be reasonably anticipated. Outputs from Stage 4 will be made available in open access peer-reviewed publications, blogs, social media and on a dedicated project website for future re-use.

**Ethical Approval:** The Institute of Population Health Research Ethics Committee of the University of Liverpool gave ethical approval for this work (REC Reference: 12413; 24th July 2023)

**Strengths and Limitations:**

- The proposed Delphi study will deliver a toolkit that assists researchers, health care organisations and policy makers decide on how to appropriately collect and use data on sensitive characteristics (e.g. sexual orientation and gender identity) including stakeholder-defined re-use of this data for specific purposes including health service improvement and developing tools for data-driven decision support (i.e. in data science, AI and ML applications designed for LGBTQI+ communities).
- This Delphi study is designed to focus on the intersection of sensitive characteristics and mental health, where similar research has focused on healthcare or sexual health more generally^1^.
- The Delphi study will be led by a team from the United Kingdom, with the expectation the consensus process will involve participants largely drawn from Western cultures with similar societal attitudes and legislative mechanisms to protect the human rights of LGBTQI+ people. This will limit the transportability and generalisability of the Delphi process and consensus outputs.

## Background

Artificial intelligence (AI), machine learning (ML) and data-driven technologies are expected to deliver novel ways of understanding and improving mental health care^2^. In healthcare applications of AI/ML generally, there has been increased focus on the potential for unintended harm arising from biases present in data^3^ and resulting from model assumptions. Two striking examples being racial biases in an algorithm deployed to identify increased healthcare needs^4^ and commonly-used models for estimating renal function (employing standard biostatistical methods) have been shown to be poorly calibrated for estimating kidney disease in people of colour^5^.

The ambition of any data-driven learning health system^6^ is to improve the care provided to patients by adapting provision to their specific needs. In the context of mental healthcare, LGBTQI+ communities are known to have specific difficulties arising from *minority stress*^7,8^ including victimisation, internalised prejudice and isolation. Consequently, LGTBQI+ people experience higher rates of suicidal distress^9^, self-harm and suicide^10^ and differential lifetime prevalence of the most common mental disorders as a function of sexual orientation and gender identity (SOGI), ethnicity and race^11^. National survey data support these studies, showing that e.g. 3% of gay and bisexual men (compared with 0.4% of men in the general UK population) attempted to end their life by suicide in 2013^12^; over 80% of trans-identifying young people have self-harmed at some point in their lives compared to around 10% in the general population^13^ and 24% had accessed mental health services^14^ in the preceding 12 months.

We note that there is variation in cultural and societal definitions of “mental health” and “mental illness”^15^, including the egregious assumption that LGBTQI+ identity is, by definition, a “mental illness”^16,17^. In this Delphi process, while we include the biomedical definition of mental illness/disorder, we will use an inclusive and broad term – “mental distress” – defined as a constellation of experiences that cause distress for the person, result in a loss of social, personal or occupational function and/or reduction in quality of life. Further, in the proposed Delphi study, mental distress is something for which the individual would seek assistance from an external source (e.g. from healthcare professionals, or peer/community support), or where other stakeholders identify an unmet need (e.g. an LGBTQI+ support community identifying lack of support for a specific set of problems in people who remain ‘invisible’ to healthcare services).

### Data Quality

Supporting LGBTQI+ people requires high-fidelity data^18,19^. However, such data is ostensibly lacking for reasons including:

- a lack of harmonisation for the recording of SOGI data resulting in fragmented, incompatible data^20,21^
- poor recording rates for local data collection, beyond services focused on, for example, cis-gendered gay men and sexual health^12^
- disclosure of SOGI characteristics to healthcare professionals is low, because LGB people experience healthcare organisations and professionals as threatening^22^ and there is evidence that an individual’s medical history, immigration status, level of internalised homophobia and degree of connectedness to the LGBTQI+ community are significant factors for disclosure with bisexual men and women being the least likely to disclose SOGI characteristics to healthcare professionals^23^
- misalignment of patient and healthcare professionals expectations around SOGI data collection, resulting in e.g. 80% healthcare professionals believing they may offend by asking about SOGI characteristics compared to 11% of patients reporting likelihood of offence^24^
- accessing healthcare is difficult for LGBTQI+ people, for example, 28% of people in the UK’s LGBT National Survey described it was “not easy” to access mental healthcare^14^

### AI, ML and Data Reuse

The straightforward imperative that we *require* better data collection is well documented^25–27^, but difficult to implement. Further, there is less evidence on the specific and acceptable uses of data and AI/ML technology to advance the provision of care for the LGBTQI+ community^28^. Therefore, there is a need to understand:

- how SOGI data can be **meaningfully collected, stored and processed** in a way that is compatible with the language and norms defined by LGBTQI+ communities
- the **current barriers to disclosure of SOGI** among LGBTQI+ people
- the **acceptable use-cases** for using individual and population level SOGI data collected in routine clinical care This paper describes a protocol for a Delphi process to develop a consensus on these three questions.

## Rationale

Patient, public and stakeholder involvement in mental health research has an established history and is motivated by^29^ stakeholder involvement as an ethical imperative with the expectation that this may improve the quality, relevance and uptake of research^30^. Arnstein’s “ladder of citizen participation”^31^ is often cited as an anchoring principle for meaningful stakeholder involvement and participatory design^32^ with contemporary definitions^33^ defining PPI as e.g. “a process whereby professionals and those traditionally on the receiving end of their ‘expertise’ (e.g. patients/service users/marginalised citizens) can collaborate with the goal of achieving outcomes that arguably cannot be achieved otherwise. It should engage the talents and experience of all involved and support the egalitarian relations and conditions needed to make the most of them”. In healthcare, the defining summary statement is “no decision about me, without me”^34,35^ and adopting this principle of empowerment and co-design for healthcare AI comes with unique challenges^36^. Participatory approaches present a necessary step in the safe development of AI systems for delivering positive impact^37^ and participatory design can play a critical role in empowering marginalised communities to take an active role in constructing research agendas and outputs; for example, in applications spanning architecture, the environment and planning^38,39^, community building^40^ and education^41^.

A central tenet of AI research applied to healthcare should be that affected communities are active participants in the co-design and production of services and technologies to avoid (usually) unintended harms, to mitigate unforeseen consequences of technical processes and the avoidance of socio-technical “blind spots”. In the application of AI specifically to LGBTQI+– inclusive mental healthcare, the interaction of minority stress^7^ with the stigmatisation of mental illness more generally^42^ presents a quagmire of acceptability, safety and healthcare equity concerns. We argue that these can only be addressed through a participatory process that identifies how services and technologies understand, collect, codify and use the communities’ data to ensure they benefit. In health sciences, the Delphi technique has been useful for establishing a consensus on “complex issues where knowledge is uncertain and incomplete”^43^ and where evidence synthesis from e.g. experimental or epidemiological data is difficult^44^. Consistent with our aims for PARQAIR-MH, the method can enable a diversity of perspectives to be represented during consensus development.

## Focus

The proposed Delphi process will focus attention on data that is expected to be collected routinely and in clinical settings (whether public, private or third-sector providers). We will not consider the re-use of data from e.g. social media sources, blogs or other self-publishing platforms. The three primary domains will be

1. How best to collect sensitive SOGI data in routine clinical practice / interactions with healthcare providers
2. Barriers/obstacles for LGBTQI+ communities including disclosure as well as people’s choices around how they access any services (public, private or third-sector) that inform why SOGI data might be systematically missing from public-sector healthcare data
3. Parameters for acceptable re-use of SOGI data beyond recording for the fidelity of an individual’s health record

Factors that explicitly address the most appropriate models of healthcare service design and delivery^14,45^ – that certainly affect people’s experiences and future engagement with providers – will be out-of-scope for PARQAIR-MH because the focus is on ways to use data to improve LGBTQI+ affirmative care.

## Methods/Design

The multistage consensus method will follow recommendations for the Delphi technique^46^ using repeated rounds of a semi-structured questionnaire with feedback from a group of stakeholders.

The Delphi process comprises multiple stages and is overseen by an executive committee. The stages are:

1. problem definition
2. selection of working groups
3. three sequential Delphi rounds, with the third round including repetition/revision of the final round alongside an estimate of consensus between participants until a pre-defined level of agreement is reached
4. transcribing, summarising and analysis of the concluding Delphi round to include final estimates of agreement/degree of consensus for the three domains
5. reporting on findings and development of consensus statement for dissemination

### Working Group

The PARticipatory Queer AI Research for Mental Health (PARQAIR-MH) working group will include:

1. an **executive committee** responsible for the overall execution of the project, organisational/operational processes to conduct and disseminate and report on the Delphi process. This group will consist of the authors of this manuscript.
2. an **advisory working group** who will lead the final-stage consensus meeting and be drawn from experts from the machine learning, ethics, health policy, mental health professionals and patient and public involvement (PPI) stakeholder groups.
3. a **survey group** of people (with similar composition to the advisory group) who will participate in the Delphi survey

The advisory and survey groups will be composed of:

1. people with lived experience of mental health service use from the LGBTQI+ communities
2. people representing charities, NGOs (non-governmental organisations) and other campaigning groups focusing on the mental health of people from the LGBTQI+ community
3. domain experts drawn from mental health professionals, invited to participate from national and international LGBTQI+ communities to include people with lived experience of mental health problems.

### Patient and Public Involvement Statement

Co-author Julia Hamer-Hunt, a lived-experience practitioner, consulted on the principles and design of the Delphi process from conception to the final draft of this protocol. The Executive Committee will be assembled by a targeted approach through the author’s professional networks, alongside open calls on social media, to ensure a diverse, equitable, inclusive and representative panel of stakeholders (including patients and public) to oversee the Working Group and execution of the Delphi consensus process.

## Stage 1: Literature review

To focus the initial round of the Delphi process, the authors of this protocol will review existing literature to identify:

- barriers to accessing mental health support implicated in the current lack of robust public sector data
- existing healthcare guidelines for the collection of SOGI data for LGBTQI+ people
- studies of perceptions, attitudes and experiences to disclosure of SOGI data in healthcare settings for LGBTQI+ people
- example applications of AI in LGBTQI+ mental health support to include those that expose benefits, risks and harms specific to that community

This review will include both published, peer-reviewed academic literature, governmental, NGO and charity surveys as well as publicly-available national policy documents. Special attention will be given to surveying the ethics and fairness literature, to identify promising approaches for ensuring privacy and safety of AI systems. The glaring absence of analyses of disparate impact of AI on queer communities^47^ further justifies the need for a deeper community involvement.

Following a review of the literature, the executive committee will produce a summary of the findings alongside a draft Terms of Reference (ToR), describing the project aims, scope and intended deliverables and outputs. This will be circulated to stakeholder groups selected by the executive committee, using their own and their organisations’ professional networks with coverage including:

- NHS and University Patient and Public Involvement/Engagement groups
- Clinicians working in the mental health sector
- Ethicists
- AI researchers and data scientists
- NGOs and charity stakeholders for LGBTQI+ mental health

The stakeholders approached to review the ToR will also be invited to join the advisory group to ensure appropriate representation.

## Stage 2: Delphi Process

### Outcome

a consensus statement that describes

1. LGBTQI+ community preferences for collecting, recording and harmonising SOGI data
2. the parameters for the acceptable (re)use of SOGI data for improving healthcare systems to include the following example use-cases:
  - the use of automation (e.g. AI-driven chatbots or recommender systems)
  - decision support (e.g. identifying risk factors for individual people)
  - configuring/commissioning services (e.g. auditing SOGI data for adapting existing, or developing new, services)
3. a checklist for AI developers to ensure a new project aligns with the needs and preferences of the LGBTQI+ community

### Design

The Delphi process will consist of three sequential rounds:

- **Round 1**: Participants will be presented with short vignettes describing either existing (from the literature review) or hypothetical use-cases for SOGI data collection and re-use. Participants will be shown questions relating to these vignettes with categorical responses and invited to provide narrative (free-text) elaborations explaining their reasoning for endorsing their answer. Thematic analysis of the narrative responses will guide generation of the round 2 survey process.
- **Round 2**: The Round 1 participants will be presented with an anonymised summary of the group’s Round 1 responses. For items in Round 1 with ≥ 70% agreement, the items will be presented as “agreed upon” and no further revisions/voting will be requested. For any additional items that emerged/were suggested, a categorical response and narrative text answer will be invited int he same format as Round 1.
- **Round 3**: A final round will aggregate responses from Round 2 to deliver a) items which have been agreed by consensus (defined as ≥ 70%) b) items that remain outstanding or contentious and could not be agreed upon and are reported as such. The resulting consensus statement will be drafted, edited and distributed to the Round 1 and 2 participants.

## Stage 3: Consensus Meeting

The output of the three Delphi rounds will be presented to a focus group for discussion and final agreement on the format of the consensus statement. Participants invited to this group will be:

- NHS and University Patient and Public Involvement/Engagement groups
- The executive committee
- Delphi rounds 1–3 participants
- Clinicians working in the mental health sector
- Ethicists
- AI researchers and data scientists
- NGOs and charity stakeholders for LGBTQI+ mental health

Importantly, we recognise that consensus may be difficult to achieve for certain topics and themes; for example, some participants might have a strong opinion that automation of any aspect of mental healthcare delivery is unacceptable. Given the complexities of defining consensus^48^, themes where consensus cannot be reached will be reported and highlighted in the final toolkit and guidance.

## Stage 4: Outcomes and Dissemination

At the consensus meeting, the outputs deemed necessary and sufficient for a toolkit will be discussed; for example, the format and medium for the researcher “checklist” and any guidance documents that would need to accompany this. It is anticipated this will take the form of a recommendations white paper and case-study format similar to e.g.^49–51^.

Following this, the executive committee will invite the advisory group to contribute to writing a summative report for submission to an open-access, peer-review journal.

The key outputs (toolkit and guidance documents for replicating the Delphi process) and findings (consensus statement including open-access, peer-reviewed papers) will be made available on a website (similar to the equator network, https://www.equator-network.org/) that will be maintained by the executive committee. The aim is to provide a participatory design-inspired open and transparent process for communities and organisations to either deploy the consensus and toolkit in their own localities, or to replicate the process to derive locally-informed versions of the toolkit/consensus.

Stakeholder involvement in all outputs from the proposed Delphi process will be transparent and explicitly described, including composition of the Executive Committee and Working Group. Specific patient and public involvement (PPI) will be reported using the GRIPP2^52^ reporting guidelines.

## Discussion

### Scope and Generality

Existing work on SOGI data collection and harmonisation reflects a largely Western geographical focus including the European Union, United Kingdom and United States^18–20^. The pending 2022 UN Report to the Human Rights Council^53^ on SOGI emphasises healthcare equity for LGBTQI+ communities (including data collection/harmonisation as a key enabler) while previous UN mandate reports^54^ acknowledge under-representation from regions of the world with hetero-normative cultural norms or where people from LGTBQ+ communities are persecuted. Similarly, different societies and cultures’ formulation of mental illness in terms of aetiology, stigma, implications for individuals, family and wider society vary to the extent that a dominantly Western biomedical model (that emphasises the individual as the locus of mental illness and disorder) is seen as unhelpful (see^15^ for a review). While the overarching PARQAIR-MH process remains general, the outcome of its initial application in the United Kingdom will be limited and localised in its immediate practical utility; necessitating replication studies.

### Limitations

The patient and public perception of clinical applications of AI is relatively under-studied; one systematic review^1^ of 23 mixed-methods studies found no studies specifically addressing mental healthcare. The review exposed some polarisation around themes of accountability (of a decision made using AI), concern around “boundary cases” (i.e. rare diseases or uncommon situations) and a divide around risk of worsening or improving healthcare outcomes, equity and justice. Importantly, they note that the perspectives of under-represented groups were rarely included or studied in the sampled literature.

Given this, we expect similar polarity in our Delphi process which may limit the extent to which consensus can be reached – given this, we will report separately on subsets of items achieving consensus, those where no consensus could be reached and a clear description of contentions arising in both subsets.

### Protocol Re-use and Utility

Considering the rising need for a wider community involvement in AI design, and this being one of the very first AI participatory studies designed specifically for the LGBTQI+ population, we are hoping that the proposed protocol will help inform a multitude of future participatory research directions. Indeed, the issues of data collection, data use, fairness and safety, are central to AI development across mental health care, healthcare, as well as numerous other domains and use cases.

Consistent with the central tenets of participatory design, this protocol needs to be applied locally, to capture the local variation in perspectives, needs, and healthcare systems. Repeated application of the protocol may result in different consensus statements, reflecting these local differences. We would therefore strongly encourage worldwide replication studies, complementing the initial study planned in the United Kingdom. In terms of utility, PARQAIR-MH aims to help inform digital health policy and the design of inclusive mental health care technologies going forward.

### Ethics and Dissemination

Participants in the Delphi process will be recruited by snowball and opportunistic sampling via professional networks and social media (but not by direct approach to healthcare service users, patients, specific clinical services or via clinicians’ caseloads). Participants will not be required to share personal narratives and experiences of healthcare or treatment for any condition. Before agreeing to participate, people will be given information about the issues considered to be in-scope for the Delphi (e.g. developing best practices and methods for collecting and harmonizing sensitive characteristics data; developing guidelines for data use/re-use) alongside specific risks of unintended harm from participating that can be reasonably anticipated. Outputs from Stage 4 will be made available in open access peer-reviewed publications, blogs, social media and on a dedicated project website for future re-use.

## Data Availability

No data has been generated in preparing this protocol.

## Authors’ Contributions

This protocol was conceived, designed and written by all five authors listed. All authors have reviewed and approved the final manuscript.

## References

1. Young, A. T., Amara, D., Bhattacharya, A. & Wei, M. L. Patient and general public attitudes towards clinical artificial intelligence: a mixed methods systematic review. The Lancet Digit. Heal. 3, e599–e611 (2021).

2. Foley, J. & Woollard, J. Digital Future of Mental Healthcare Report. https://topol.hee.nhs.uk/wp-content/uploads/HEE-Topol-Review-Mental-health-paper.pdf (2019).

3. Chen, R. J. et al. Algorithm fairness in AI for medicine and healthcare. arXiv preprint arXiv:2110.00603 DOI: https://doi.org/10.48550/arXiv.2110.00603 (2021).

4. Obermeyer, Z., Powers, B., Vogeli, C. & Mullainathan, S. Dissecting racial bias in an algorithm used to manage the health of populations. Science 366, 447–453 (2019).

5. Diao, J. A., Powe, N. R. & Manrai, A. K. Race-Free Equations for eGFR: Comparing Effects on CKD Classification. J. Am. Soc. Nephrol. 32, 1868–1870 (2021).

6. McLachlan, S. et al. The Heimdall framework for supporting characterisation of learning health systems. BMJ Heal. & Care Informatics 25, 77–87, DOI: 10.14236/jhi.v25i2.996 (2018).

7. Meyer, I. H. Prejudice, social stress, and mental health in lesbian, gay, and bisexual populations: Conceptual issues and research evidence. Psychol. Bull. 129, 674–697, DOI: 10.1037/0033-2909.129.5.674 (2003).

8. Hendricks, M. L. & Testa, R. J. A conceptual framework for clinical work with transgender and gender nonconforming clients: An adaptation of the Minority Stress Model. Prof. Psychol. Res. Pract. 43, 460–467, DOI: 10.1037/a0029597 (2012).

9. Johnson, K., Faulkner, P., Jones, H. & Welsh, E. Understanding Suicidal Distress and Promoting Survival in Lesbian, Gay, Bisexual and Transgender (LGBT) Communities. https://citeseerx.ist.psu.edu/viewdoc/download?doi=10.1.1.627.8017&rep=rep1&type=pdf (2007).

10. Williams, A. J. et al. A systematic review and meta-analysis of victimisation and mental health prevalence among LGBTQ+ young people with experiences of self-harm and suicide. PLOS ONE 16, e0245268, DOI: 10.1371/journal.pone.0245268 (2021).

11. Meyer, I. H., Dietrich, J. & Schwartz, S. Lifetime Prevalence of Mental Disorders and Suicide Attempts in Diverse Lesbian, Gay, and Bisexual Populations. Am. J. Public Heal. 98, 1004–1006, DOI: 10.2105/AJPH.2006.096826 (2008).

12. Women and Equalities Committee. Health and Social Care and LGBT Communities. https://publications.parliament.uk/pa/cm201919/cmselect/cmwomeq/94/94.pdf (2019).

13. Stonewall. LGBT in Britain – Health. https://www.stonewall.org.uk/lgbt-britain-health (2018).

14. Government Equalities Office. National LGBT Survey: Research report. https://www.gov.uk/government/publications/national-lgbt-survey-summary-report (2018).

15. Gopalkrishnan, N. Cultural diversity and mental health: Considerations for policy and practice. Front. Public Heal. 6, 179 (2018).

16. King, M., Smith, G. & Bartlett, A. Treatments of homosexuality in britain since the 1950s—an oral history: the experience of professionals. BMJ 328, 429 (2004).

17. King, M. Stigma in psychiatry seen through the lens of sexuality and gender. BJPsych Int. 16, 77–80 (2019).

18. HRC.org. LGBTQ-Inclusive Data Collection: A Lifesaving Imperative. https://assets2.hrc.org/files/assets/resources/HRC-LGBTQ-DataCollection-Report.pdf (2019).

19. Bell, M. Data collection in relation to LGBTI People. https://ec.europa.eu/newsroom/just/redirection/document/45605 (2017).

20. Office for National Statistics. Equalities data audit (final report). https://www.ons.gov.uk/methodology/methodologicalpublications/generalmethodology/onsworkingpaperseries/equalitiesdataauditfinalreport (2018).

21. Young, W. J., Bover Manderski, M. T., Ganz, O., Delnevo, C. D. & Hrywna, M. Examining the impact of question construction on reporting of sexual identity: Survey experiment among young adults. JMIR Public Heal. Surveill 7, e32294, DOI: 10.2196/32294 (2021).

22. Eliason, M. J. & Schope, R. Does “Don’t Ask Don’t Tell” Apply to Health Care? Lesbian, Gay, and Bisexual People’s Disclosure to Health Care Providers. J. Gay Lesbian Med. Assoc. 5, 125–134, DOI: 10.1023/A:1014257910462 (2001).

23. Durso, L. E. & Meyer, I. H. Patterns and Predictors of Disclosure of Sexual Orientation to Healthcare Providers Among Lesbians, Gay Men, and Bisexuals. Sex. Res. Soc. Policy 10, 35–42, DOI: 10.1007/s13178-012-0105-2 (2013).

24. Maragh-Bass, A. C. et al. Risks, Benefits, and Importance of Collecting Sexual Orientation and Gender Identity Data in Healthcare Settings: A Multi-Method Analysis of Patient and Provider Perspectives. LGBT Heal. 4, 141–152, DOI: 10.1089/lgbt.2016.0107 (2017).

25. Grasso, C., Goldhammer, H., Thompson, J. & Keuroghlian, A. S. Optimizing gender-affirming medical care through anatomical inventories, clinical decision support, and population health management in electronic health record systems. J. Am. Med. Informatics Assoc. 28, 2531–2535, DOI: 10.1093/jamia/ocab080 (2021).

26. Keuroghlian, A. S. Electronic health records as an equity tool for LGBTQIA+ people. Nat. Medicine 1–3, DOI: 10.1038/s41591-021-01592-3 (2021).

27. National LGBT Cancer Network. Ending the Invisibility: Organizations Call for Routine LGBTQI+ Data Collection. https://cancer-network.org/ending-the-invisibility-organizations-call-for-routine-lgbtqi-data-collection/ (2021).

28. Tomasev, N., McKee, K. R., Kay, J. & Mohamed, S. Fairness for Unobserved Characteristics: Insights from Technological Impacts on Queer Communities. Proc. 2021 AAAI/ACM Conf. on AI, Ethics, Soc. 254–265, DOI: 10.1145/3461702.3462540 (2021).

29. Boivin, A. et al. Evaluating patient and public involvement in research. BMJ 363, k5147 (2018).

30. O’Brien, J., Fossey, E. & Palmer, V. J. A scoping review of the use of co-design methods with culturally and linguistically diverse communities to improve or adapt mental health services. Heal. & Soc. Care Community 29, 1–17 (2021).

31. Arnstein, S. R. A ladder of citizen participation. J. Am. Inst. planners 35, 216–224 (1969).

32. Szebeko, D. & Tan, L. Co-designing for society. Australas. Med. J. 3 (2010).

33. Ocloo, J., Garfield, S., Franklin, B. D. & Dawson, S. Exploring the theory, barriers and enablers for patient and public involvement across health, social care and patient safety: a systematic review of reviews. Heal. research policy systems 19, 1–21 (2021).

34. Coulter, A. & Collins, A. Making shared decision-making a reality. https://ugc.futurelearn.com/uploads/files/19/40/19408460-e688-4a99-84bb-d5114eca9c97/2.3_Making-shared-decision-making-a-reality-paper-Angela-Coulter-Alf-Collins-July-2011_0.pdf (2011).

35. Slade, M. Implementing shared decision making in routine mental health care. World Psychiatry 16, 146–153 (2017).

36. Donia, J. & Shaw, J. A. Co-design and ethical artificial intelligence for health: An agenda for critical research and practice. Big Data & Soc. 8, 1–12 (2021).

37. Bondi, E., Xu, L., Acosta-Navas, D. & Killian, J. A. Envisioning communities: a participatory approach towards AI for social good. In Proceedings of the 2021 AAAI/ACM Conference on AI, Ethics, and Society, 425–436 (2021).

38. Broto, V. C. Queering participatory planning. Environ. Urbanization 33, 310–329 (2021).

39. Hardy, J. & Vargas, S. Participatory design and the future of rural LGBTQ communities. In Companion Publication of the 2019 on Designing Interactive Systems Conference 2019, 195–199, DOI: https://doi.org/10.1145/3301019.3323894 (2019).

40. Oaks, L., Israel, T., Conover, K. J., Cogger, A. & Avellar, T. R. Community-based participatory research with invisible, geographically-dispersed communities: Partnering with lesbian, gay, bisexual, transgender and queer communities on the California central coast. J. for Soc. Action Couns. & Psychol. 11, 14–32 (2019).

41. McWilliams, J. Queering participatory design research. Cogn. Instr. 34, 259–274 (2016).

42. Crisp, A. H., Gelder, M. G., Rix, S., Meltzer, H. I. & Rowlands, O. J. Stigmatisation of people with mental illnesses. The British Journal of Psychiatry 177, 4–7 (2000).

43. Niederberger, M. & Spranger, J. Delphi technique in health sciences: a map. Front. public health 8, 457 (2020).

44. Jorm, A. F. Using the delphi expert consensus method in mental health research. Aust. & New Zealand J. Psychiatry 49, 887–897 (2015).

45. Pachankis, J. E., Clark, K. A., Jackson, S. D., Pereira, K. & Levine, D. Current capacity and future implementation of mental health services in US LGBTQ community centers. Psychiatr. services 72, 669–676 (2021).

46. Linstone, H. A. & Turoff, M. The Delphi Method: Techniques and Applications (Addison-Wesley Reading, MA, 1975).

47. Birhane, A. et al. The Forgotten Margins of AI Ethics. In 2022 ACM Conference on Fairness, Accountability, and Transparency, FAccT ‘22, 948–958, DOI: 10.1145/3531146.3533157 (Association for Computing Machinery, New York, NY, USA, 2022).

48. Diamond, I. R. et al. Defining consensus: a systematic review recommends methodologic criteria for reporting of Delphi studies. J. Clin. Epidemiol. 67, 401–409 (2014).

49. Ada Lovelace Institute. Participatory data stewardship. https://www.adalovelaceinstitute.org/report/participatory-data-stewardship/ (2021).

50. Ada Lovelace Institute. Algorithmic impact assessment: a case study in healthcare. https://www.adalovelaceinstitute.org/ wp-content/uploads/2022/02/Algorithmic-impact-assessment-a-case-study-in-healthcare.pdf (2022).

51. Reisman, D., Schultz, J., Crawford, K. & Whittaker, M. Algorithmic impact assessments: a practical framework for public agency accountability. https://ainowinstitute.org/aiareport2018.pdf (2018).

52. Staniszewska, S. et al. GRIPP2 reporting checklists: tools to improve reporting of patient and public involvement in research. BMJ 358 (2017).

53. UNHRC. Call for inputs: Report to the UN Human Rights Council on the realisation of the right of persons affected by violence and discrimination based on sexual orientation and gender identity to the enjoyment of the highest attainable standard of physical and mental health, in relation to SDG3. https://www.ohchr.org/en/calls-for-input/calls-input/call-inputs-report-un-human-rights-council-realisation-right-persons (2022).

54. UNHRC. Protection against violence and discrimination based on sexual orientation and gender identity. https://documents-dds-ny.un.org/doc/UNDOC/GEN/N21/192/14/PDF/N2119214.pdf?OpenElement (2021).

